# Impact of temporary storage conditions on the viability of *Streptococcus pneumoniae* in saliva

**DOI:** 10.1101/2022.06.27.22276954

**Authors:** Orchid M. Allicock, Anna York, Pari Waghela, Devyn Yolda-Carr, Daniel M. Weinberger, Anne L. Wyllie

## Abstract

Nasopharyngeal swabs are considered the gold standard sample type for the detection of *Streptococcus pneumoniae* carriage, but recent studies have demonstrated the utility of saliva in improving carriage detection in adults. Saliva is generally collected in its raw, unsupplemented state, unlike nasopharyngeal swabs, which are collected into stabilizing transport media. Little data exist regarding the stability of pneumococcus in unsupplemented saliva during transport and laboratory storage. We therefore evaluated the effect of storage conditions on the detection of pneumococcus in saliva samples using strains representing eight pneumococcal serotypes. The bacteria were spiked into raw saliva from asymptomatic individuals, and we assessed sample viability after storage for up to 72 hours at 4°C and room temperature as well as following three freeze-thaw cycles. We observed little decrease in pneumococcal detection following culture-enrichment and qPCR detection of genes *piaB* and *lytA* when compared to testing fresh samples, indicating prolonged viability of pneumococcus in neat saliva samples. This study provides insight into the effect of storage of saliva samples in the laboratory, and the utilization of saliva for pneumococcal carriage detection which can be particularly useful for studies conducted in remote or low resource settings.

## Introduction

*Streptococcus pneumoniae* (pneumococcus) is among the most frequent causes of severe pneumonia and pneumonia deaths in the United States (1). However, pneumococcus is also a commensal organism that resides asymptomatically in the upper respiratory tract of healthy individuals. In susceptible individuals, pneumococcus can become pathogenic under certain circumstances. Pneumococcus is the causative agent of otitis media, sinusitis and pneumonia as well as invasive pneumococcal diseases (IPD), such as meningitis and sepsis. The rate of morbidity and mortality for serious pneumococcal disease is highest in young children, older adults, and immunocompromised individuals (2). Case fatality rates range up to 20% for sepsis and 50% for meningitis in lower and middle income countries (3). Following the introduction of pneumococcal conjugate vaccines (PCV) 7 and 13 (PCV7 and PCV13), both asymptomatic carriage and IPD caused by vaccine serotypes declined. However, there was a concomitant increase in the prevalence of non-vaccine serotypes (NVTs), known as serotype replacement (4-6). Post-vaccine studies have provided insights into the shift in frequency of vaccine and non-vaccine serotypes in both carriage and disease (7-10). However, continued large-scale surveillance is needed to monitor the changing serotype landscape in order to better inform the development of next-generation vaccines.

Nasopharyngeal swabs have long been considered the gold-standard sample type for detection of carriage of pneumococcus in young children, however detecting carriage in adults is more challenging. In 2013, the WHO advised the additional collection of oropharyngeal swabs in older adults (11), although false-positivity from other oral streptococci may be problematic (12). Due to their invasive nature, nasopharyngeal swabs are generally not well tolerated, and sample collection requires a healthcare worker. This can restrict or prevent sampling of specific populations, particularly the elderly or disabled or individuals in low-resource settings with minimal access to healthcare. Therefore, alternative, less-invasive sample types, which can be collected without the need for healthcare workers, are important for the study of pneumococcal carriage in these populations.

The use of nasopharyngeal swabs for pneumococcal carriage studies during the COVID-19 pandemic was associated with safety and supply chain issues. Non-essential in-person visits with healthcare workers were restricted due to safety concerns. Additional supply chain issues reduced access to materials required for nasopharyngeal and oropharyngeal sampling, further hampering attempts to continue carriage studies. These complications provided the impetus for pneumococcal carriage studies to move away from swab-based sampling and utilize alternative sample types. Saliva has been shown to be a sensitive sample type for detection of pneumococcal carriage in individuals of all ages (13-15). Saliva sampling is non-invasive and can be collected quickly and easily in the home without a healthcare worker. However, at-home sample collection brings about potential issues with short-term storage, collection and transport of the samples to the lab. In order to reap the benefits of using saliva samples for studies of pneumococcal carriage, more data are needed about how variations in storage conditions and time from sample collection to laboratory processing may impact detection and viability of pneumococcus in these samples.

In this study, we evaluated the viability of pneumococcus in saliva over extended periods (24, 48 and 72 hours) when stored at either 4°C or room temperature. We also investigated the viability of pneumococcus after a series of freeze-thaw cycles at -20°C and -80°C, with and without glycerol supplementation to evaluate best practices for storage and processing of saliva samples in the laboratory. Understanding the stability of pneumococcus in saliva is also particularly important for carriage studies being conducted in remote regions or in resource-limited settings where quick transport or access to cold storage may not be readily available.

## Methods

### Ethics statement

Deidentified saliva samples were collected from healthy adult volunteers who were asymptomatic for respiratory infection. Potential study participants were informed in writing about the purpose and procedure of the study and consented to study participation through the act of providing a saliva sample; the requirement for written informed consent was waived by the Institutional Review Board of the Yale Human Research Protection Program (HIC #2000029374).

### Saliva samples

Saliva samples were collected from healthy volunteers according to previously published protocols (15). Participants were asked to drool saliva into a 15 mL polypropylene tube at least one hour after their last meal or drink, these saliva samples were considered whole mouth unstimulated saliva. Samples were transferred at room temperature to the laboratory for temporary storage at 4°C and stored within 12 hours at -80°C.

### Sample processing

To confirm the status of each saliva sample (i.e. pneumococcus positive or negative), 50 µl of saliva was heated for 10 mins at 95°C, and a qPCR targeting *piaB* and *lytA* was conducted (as described below). Saliva samples that were qPCR-negative for *piaB* were considered pneumococcal negative and were stored at -20°C until required for spiking experiments.

### Bacterial Isolates

Pneumococcal isolates from our collection (Table 1) were selected to represent eight different representative serotypes (3, 4, 5, 6C, 7F, 9V, 11A, 19F). These isolates were cultured on TSA II + 5% (v/v) defibrinated sheep blood (blood plates) and incubated at 37°C with 5% CO_2_ overnight. Serotypes were confirmed using IMMULEX™ pneumococcus antisera per the manurfacturer’s instructions. The isolates’ lawn was harvested into brain heart infusion (BHI) media supplemented with 10% (v/v) glycerol (BHI+10% glycerol) using a cotton swab and was stored at -80°C for spiking. The colony forming units per mL (CFU/mL) of each sample was determined by colony counting of serially diluted samples, cultured on blood plates and incubated at 37°C and 5 % CO_2_ overnight.

**Table 1.** Strains of *Streptococcus pneumoniae* used in this study.

| Strain | Serotype | Source |
| --- | --- | --- |
| ABC020026160 | 3 | CDC |
| ABC020012001 | 4 | CDC |
| ABC020024416 | 5 | CDC |
| ABC020025076 | 6C | CDC |
| ABC010005954 | 7F | CDC |
| ABC010001831 | 9V | CDC |
| ABC020009080 | 11A | CDC |
| #64Israel | 19F | (16) |

### Experimental Design

For each isolate listed in **Table 1**, the spiked-saliva experiments were conducted at 10^3^ and 10^4^ CFU/mL which are clinically relevant for saliva samples in children (Wyllie et al. 2014; York et al. 2022). To determine the stability of pneumococcus over time, spiked and clinical saliva samples were incubated at room temperature (∼20°C) or 4°C. At 24 hours, 48 hours and 72 hours, the spiked-saliva samples were vortexed, and 100 µl were removed from each sample for culture-enrichment.

To determine the effect of freeze-thawing on the stability of pneumococcus, aliquots of spiked-saliva (10^3^ and 10^4^ CFU/mL), with and without the addition of 10% glycerol, were stored for a minimum of 2 hours at -20°C or -80°C, before being thawed to room temperature. Once each of the aliquots was thawed, the samples were vortexed, and 100 µl were removed from each sample for culture-enrichment before the remainder was returned to -20°C or -80°C. This process was repeated twice more.

### Culture enrichment of saliva

At each of the study time points, 100 µl of spiked saliva or clinical saliva was plated on TSA II + 5% (v/v) defibrinated sheep blood supplemented with 10 µg/mL gentamicin (gent plates) (Krone et al. 2015), and incubated at 37°C and 5% CO_2_ overnight. The following day, all bacterial growth was harvested from culture plates into BHI+10% glycerol, and stored at -80°C until further analysis. These stored samples were considered to be culture-enriched for pneumococci. Culture-enrichment of saliva samples at each time-point ensures that Ct values obtained represent viable pneumococci and not exogenous DNA.

### Detection of pneumococcal carriage using the molecular method

Culture-enriched saliva samples were thawed on ice. DNA was extracted from 200 μl of each sample using the MagMAX Ultra Viral/Pathogen Nucleic Acid Isolation kit (ThermoFisher Scientific) on the KingFisher Apex (ThermoFisher Scientific) with modifications. Each DNA template was tested in qPCR for pneumococcal genes *piaB (17, 18)* and *lytA* (19). The assays were carried out in 20 µl reaction volumes using SsoAdvanced Universal Probe Supermix (Biorad, USA), primer/probe mixes (Iowa Black quenchers) for *piaB* and *lytA* at final concentrations of 10 nM and 12 nM respectively, and 2.5 µl of genomic DNA. DNA of *S. pneumoniae* serotype 19F strain was included as a positive control in every run. Assays were run on a CFX96 Touch (Biorad) under the following conditions: 95°C for 3 minutes, followed by 45 cycles of 98°C for 15 seconds and 60°C for 30 seconds. Samples were considered positive for pneumococcus when Ct values for both genes were ≤40 (13).

### Colony isolation and serotyping

Clinical samples that were qPCR-positive for *piaB* (pneumococcus-positive), were serially diluted in phosphate buffered saline, then 100 µl of 10^−4^, 10^−5^ and 10^−6^ dilutions were plated on blood plates, and incubated at 37°C and 5% CO_2_. Following overnight incubation, plates were screened for single alpha-haemolytic colonies, which were selected, expanded onto new plates, and tested for optochin sensitivity. Optochin sensitive isolates were serotyped using IMMULEX™ pneumococcus antisera (SSI Diagnostica, Hillerød, Denmark) per the manufacturer’s instructions.

### Statistical analysis

To evaluate the impact of temperature, serotype, number of freeze-thaw cycles and concentration of spiked bacteria on the recovery of pneumococcus from spiked saliva samples, linear regression was used. Interaction terms were used to evaluate whether the effect of time, temperature, or starting concentration varied by strain. ΔCt represented the change in Ct value from freshly spiked saliva following each condition (categorical). P-values of less than 0.05 were considered significant. Statistical tests were conducted using R (version 3.5.2) as described in the figure legends.

## Results

The viability of pneumococcus in a sample was inferred by the detection of pneumococcal DNA after undergoing sample culture enrichment. Overall, the Ct values obtained for both gene targets *piaB* and *lytA* were concordant (see Tables S1 - S3). For clarity only *piaB* results will be presented and discussed.

### Viability of *S. pneumoniae* over time

To examine the viability of *S. pneumoniae* in raw saliva samples at temporary storage conditions, we measured the concentration of pneumococcal DNA following culture enrichment by qPCR. When comparing two concentrations of spiked saliva (10^3^ and 10^4^ CFU/mL), averaging across all serotypes at time zero, Ct values of spiked saliva at 10^3^ CFU/mL were on average 2.29 higher than those at 10^4^ CFU/mL (*p*<0.001) (**Figure 1**). After adjusting for concentration, there was no significant increase in Ct (which would represent a decline in bacterial survival) between 0 and 24 hours (ΔCt 1.08, *p*=0.12), however after 48 and 72 hours a significant increase in Ct value was observed (ΔCt 3.11, *p*<0.001 and ΔCt 3.18, *p*<0.001 respectively). The pneumococci in saliva samples were slightly more stable at 4°C, but this was not significant (ΔCt 0.17, *p*=0.73).

**Figure 1.**
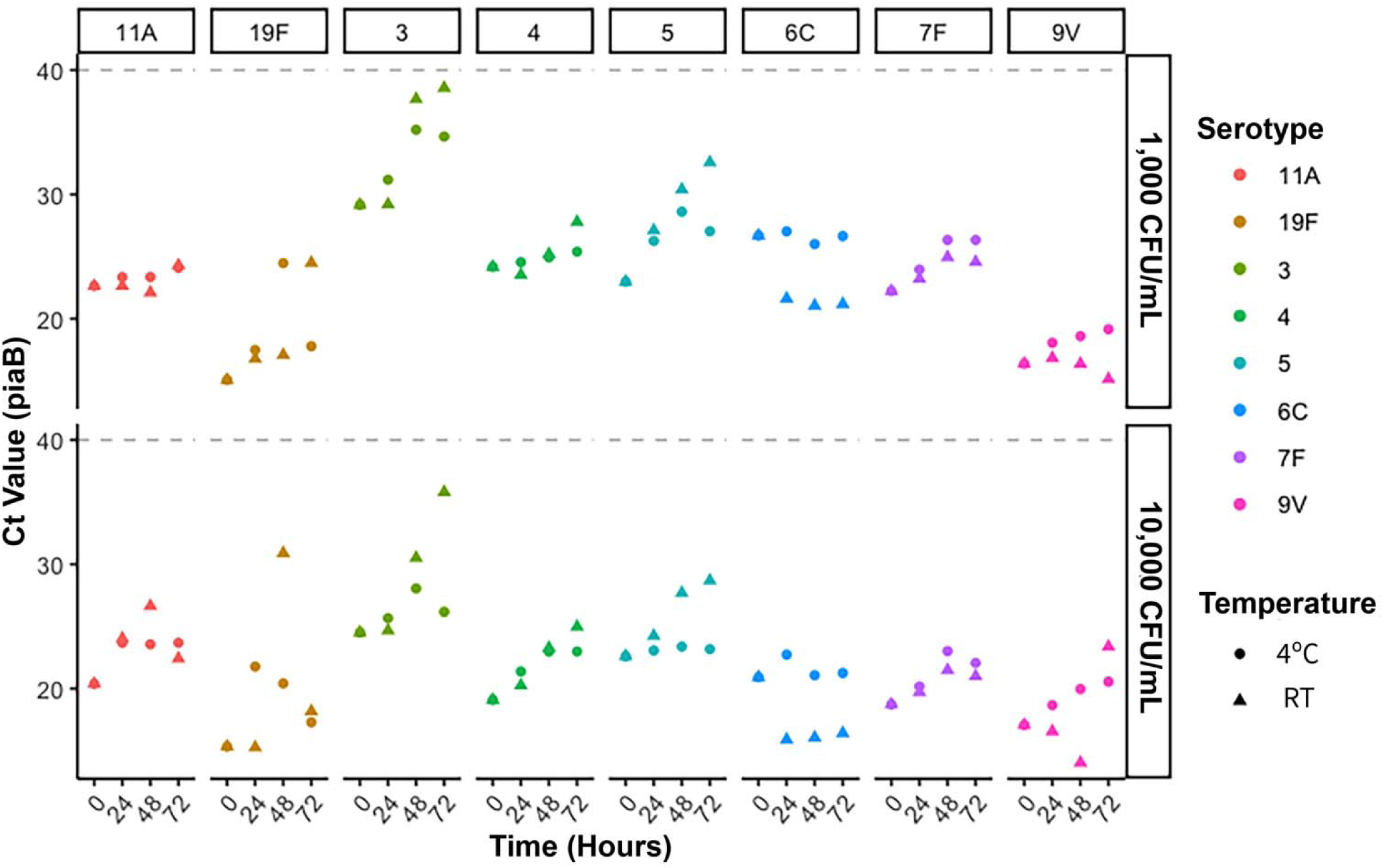
Viability of *Streptococcus pneumoniae* after prolonged storage at room temperature or 4°C. *S. pneumoniae* detection in saliva spiked at 1,000 CFU/mL and 10,000 CFU/mL over a 72-hour time course when stored at either 4°C (circles) or room temperature (triangles). Detection of strains representing each serotype was relatively consistent for up to at least 48 hours. Each point represents an individual sample. *lytA* values can be found in Table S1.

### Viability of *Streptococcus pneumoniae* after freeze-thaw cycles

Saliva from healthy volunteers were spiked with *S. pneumoniae* at different concentrations, and after each freeze-thaw cycle, we measured pneumococcus from all the strains tested, representing eight different serotypes. At both conditions (−20°C or -80°C) there was no significant difference in Ct values between fresh saliva and the first freeze-thaw cycle (**Figure 2)**. However, pneumococcal loss was lower at -80°C than at -20°C for both cycle 2 (−80°C, ΔCt -3.81 *p*=0.01 vs -20°C, ΔCt 3.877, *p*<0.001) and cycle 3 (−80°C, ΔCt -4.65, *p*<0.05 vs -20°C, ΔCt 5.056, *p*<0.001) when compared to fresh saliva. The addition of glycerol to the saliva samples before storage as a cryoprotectant did not make an impact on survival at any stage, irrespective of temperature (*p=*0.1). Of the 8 strains tested, the increase in Ct value was most notable for the isolates with serotype 3, 4 and 6C capsules, where no viable pneumococcus was detected after cycle 2, 3 or 2 respectively.

**Figure 2.**
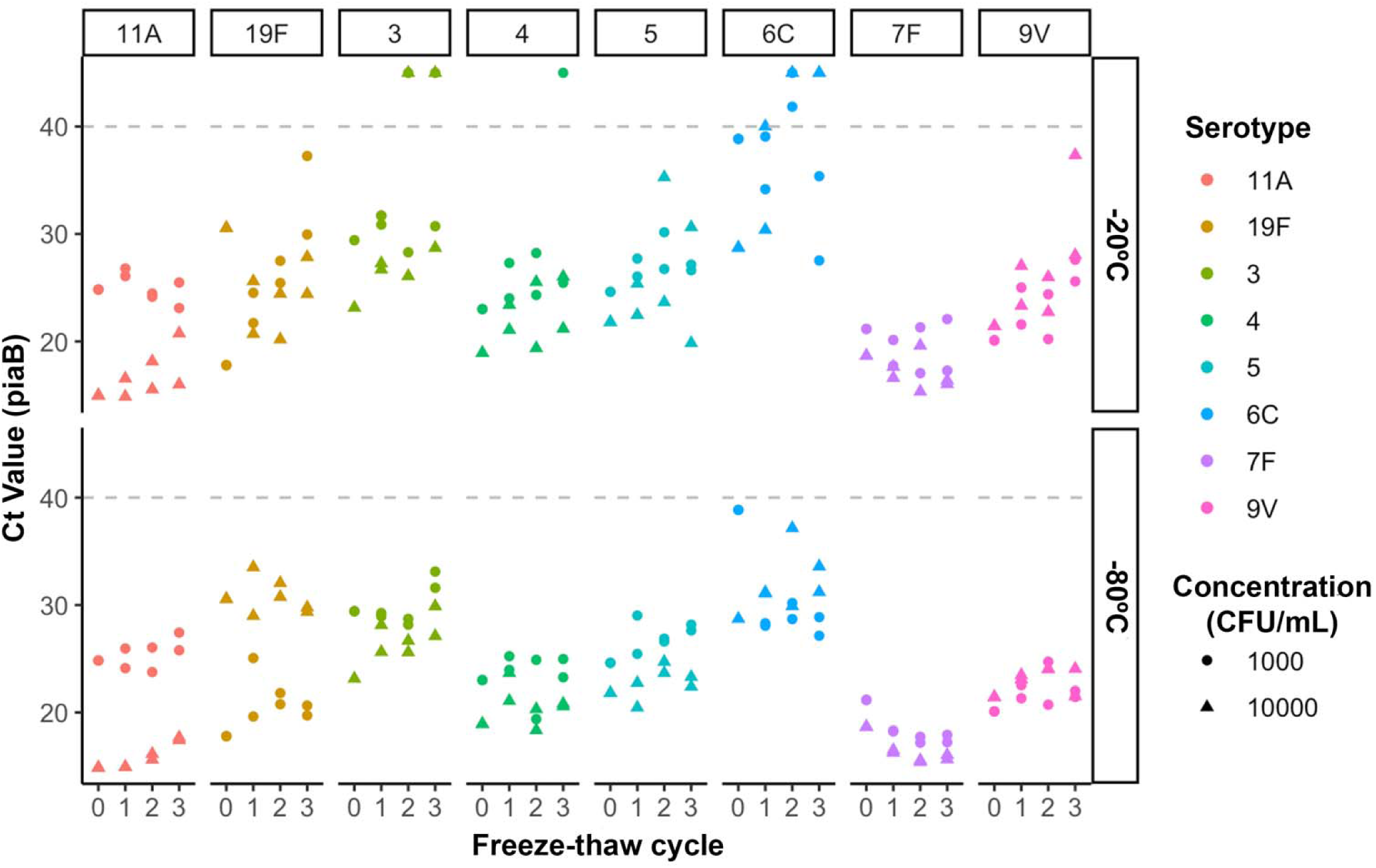
Viability of *Streptococcus pneumoniae* after freeze-thaw cycles. *S. pneumoniae* detection in saliva spiked at 1,000 CFU/mL (circles) and 10,000 CFU/mL (triangles) following each over a series of three freeze-thaw cycles at either -20°C or -80°C. *lytA* values can be found in Table S2.

### Clinical validation

Pneumococcus was detected in the saliva specimens collected from three asymptomatic volunteers (referred to as P1, P2 and P3). These saliva specimens were aliquoted and subjected to the conditions outlined previously. The viability of pneumococcus in specimens P1 and P2 (Ct values when tested fresh of 33.04, 19.65 respectively) remained stable over three freeze-thaw cycles and at both 4°C and room temperature over 72 hours (**Figure 3**). However, for specimen P3 (Ct values when tested fresh of 36.99), pneumococcus was only detected after the first freeze-thaw cycle, but not thereafter. Detection remained stable at 72 hours at room temperature, but for only 48 hours at 4°C. Notably, there was discordance between Ct values for *piaB* and *lytA* (difference of >5 Ct values) in both between *piaB* and *lytA* genes for P2 and P3 specimens, suggesting possible carriage of non-pneumococcal streptococci with a *lytA* homologue (20-22) or potentially unencapsulated pneumococci (23).

**Figure 3.**
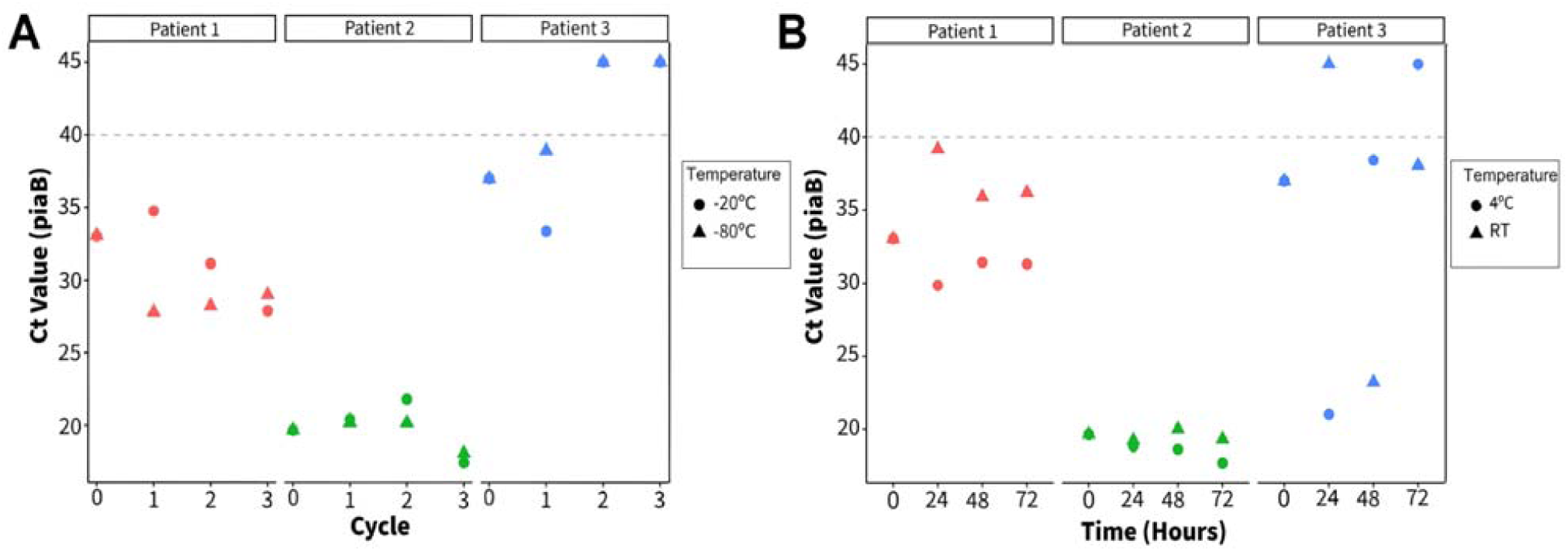
Detection of *S. pneumoniae* in clinical samples (P1-3) over **A** three freeze-thaw cycles, and **B** over a 72-hour time course. We were able to isolate multiple pneumococcal colonies from P2, however we had difficulty isolating colonies from P1 and P3. Abbreviations: Cy – cycles; H – hours; RT – room temperature. *lytA* values can be found in Table S3.

An attempt was made at isolating the pneumococcal colonies. For P2, of the 36 colonies that were optochin sensitive, 32 were identified to be the majority species belonging to serogroup 4, and 4/36 were identified to be a minority co-colonizing species belonging to serogroup 15. We were unable to isolate single colonies of pneumococcus from the other clinical samples using standard culture-based methods.

## Discussion

New PCVs that target different sets of serotypes are in various stages of clinical development and introduction in the community. Ongoing surveillance of pneumococcus in the community is necessary to assess the impact of PCVs and to monitor the emergence of non-vaccine serotypes. Nasopharyngeal swabs and/or oropharyngeal swabs are considered the gold standard sample type for surveillance, however they have their limitations. They are not well tolerated due to the discomfort, resulting in many individuals showing reluctance or poor adherence to sampling protocols. The requirement for a healthcare professional to collect the sample can cause additional challenges. The COVID-19 pandemic meant that collection of nasopharyngeal swabs for surveillance studies was difficult, and alternative sample types were needed urgently. While collection of saliva is sensitive for detection of pneumococcal carriage (14, 15), the viability of pneumococcus in saliva was previously unknown.

For this study, we selected eight strains, representing eight different serotypes (3, 4, 5, 6C, 7F, 9V, 11A and 19F) with varying degrees of encapsulation and different capsular properties. Serotypes 4, 11A and 19F represent small, medium and large capsules respectively (24). Serotype 3 was included in this study due to its clinical significance; it often causes severe disease, and is able to evade the vaccine induced immune response by releasing its capsule into the surrounding environment (25). While the majority of serotypes are anionic, serotype 7F (along with 7A, 14, 33F, 33A, and 37) is uncharged (26, 27) and was therefore also included in this study. As *piaB* was used to determine the presence of pneumococci in spiked-saliva, and piaB is absent from the majority of unencapsulated pneumococci (23), we did not include any unencapsulated isolates in this study. Any future work including the viability of unencapsulated pneumococci may require alternate gene targets such as SP2020 (22).

For the eight isolates tested, we demonstrated that saliva samples can be stored or transported at room temperature or 4°C for up to 48 hours (and often longer) without any significant loss of viability to pneumococcus. All strains of the eight serotypes tested showed no significant loss in survival over the first 24 hours. Between 24 and 72 hours some decrease in survival was observed, representing a 10-fold loss in bacterial viability, but all strains were still detected regardless of starting concentration. Therefore, for the storage and transport of saliva collected for pneumococcal carriage studies, either room-temperature or refrigeration are feasible.

In contrast, for some strains, multiple freeze-thaw cycles resulted in a decrease or loss of detection, representing to us an effect on strain viability. Furthermore, we observed no benefit by use of 10% glycerol as a cryoprotectant in this sample type. For long-term laboratory storage, samples should be aliquoted to minimize the number of freeze-thaw cycles that occur.

Differences in isolation of pneumococci from the three clinical samples (P1, P2 and P3) were related to the concentration of pneumococci in the saliva. The Ct value of P2 suggests that the pneumococcal concentration was between 10^3^ and 10^4^ CFU/mL whereas both P1 and P3 contain less than 10^1^ CFU/mL. Using culture-based methods, pneumococci could easily be isolated from sample P2 due to its high concentration. Additional enrichment steps, such as Magnetic Bead-Based Separation may be required for isolation of pneumococci from P1 and P3 due to the reduced pneumococcal concentration and the high concentration of other bacterial species in saliva (reflected in the high Ct values) (28).

For the first time, this study investigates the viability of pneumococci in raw-unsupplemented saliva at temporary storage conditions, supporting the use of saliva as a sample type for pneumococcal carriage. With little difference observed between the strains tested, findings suggest that pneumococcal survival in saliva may be independent of capsule size and charge. However, since each serotype is represented by only one isolate, additional studies are recommended to further investigate this observation at the serotype level. The findings presented here are particularly important for studies conducted in low-resource settings, rural areas and for studies conducted in the absence of healthcare professionals and adequate cold-chain transfer of samples between sampling and processing sites. In addition to being cost effective, and easy for self-collection, the use of saliva for pneumococcal carriage studies may increase participant enrollment and retention, which is particularly important for longitudinal studies. The impact of storage conditions may vary between host saliva and so our work could be extended to samples from a larger subject population, with differing saliva microbial communities.

## Data Availability

All data produced in the present work are contained in the manuscript

